# Overcoming the pitfalls of NGS-based molecular diagnosis of Shwachman-Diamond syndrome

**DOI:** 10.1101/2021.08.22.21262444

**Authors:** Xiaomin Peng, Xinran Dong, Yaqiong Wang, Bingbing Wu, Huijun Wang, Wei Lu, Feifan Xiao, Lin Yang, Gang Li, Wenhao Zhou, Bo Liu, Yulan Lu

## Abstract

**Purpose:** Shwachman-Diamond syndrome (SDS) is predominately caused by biallelic mutations in the *SBDS* gene and is characterized by exocrine pancreatic insufficiency, skeletal abnormalities and pancytopenia. Gene conversion between *SBDS* and its pseudogene *SBDSP1* is the major cause. We established an efficient approach, HapICE, to infer the haplotype of *SBDS* based on short-read next-generation sequencing (NGS).

**Methods:** HapICE based on the Expectation-Maximization algorithm was developed to detect variants in exon 2 of *SBDS* arising from gene conversion. We retrospectively analyzed two common pathogenic variants (c.183_184delinsCT and c.258+2T>C) in suspected SDS patients and compared the results with those from conventional NGS analysis.

**Results:** In 47 SDS high-risk individuals and 64 available parents, HapICE improved the diagnostic rate by 27.7% compared with conventional methods and revealed 100% (95% CI: 92.5%-100%) concordance with Sanger sequencing. In addition to eighteen patients having consistent genetic results by both methods, HapICE further reported 8 patients with more accurate variant detection and 13 cases with the c.183_184delinsCT variant missing by conventional methods. HapICE also showed better performance in screening for carrier and wild-type status.

**Conclusion:** We have developed a novel *SBDS* variant detection tool through regular NGS data that demonstrated precise variant detection performance in clinical scenarios.

## 1. Introduction

Short-read next-generation sequencing (NGS) has revolutionized the molecular diagnosis of genetic diseases and become the first-tier diagnostic assay in many clinical practices ^[1-4]^. However, researchers and clinicians continue to face challenges due to high sequence similarity between homologous genes/regions and the accompanying gene conversion events. Gene conversion, a homologous recombination mechanism, often occurs between highly homologous parental genes/pseudogenes sharing a sequence similarity ranging from 92–99% ^[5, 6]^. It is generally involved in the substitution of at least two neighboring but nonconsecutive markers within a short sequence^[5]^. To date, abundant gene conversion events have been implicated as molecular causes and they play vital roles in human genetic disorders ^[7-10]^. However, data analysis derived from short-read NGS is often hindered by pseudogene loci that are almost identical to orthologous loci and prevent the unique alignment of sequencing reads. Ambiguous read mapping will lead to inaccurate variant detection and concomitant misdiagnosis issues. Thus, how to accurately detect pathogenic variants resulting from gene conversion in homologous regions remains a challenge for NGS data analysis.

Shwachman-Diamond syndrome (SDS, MIM: 260400) is an autosomal recessive inherited disorder characterized by skeletal dysplasia, exocrine pancreatic dysfunction and pancytopenia ^[11]^. It is the second most common cause of exocrine pancreatic insufficiency, with a reported incidence of 1 in 75,000 individuals ^[12]^. Biallelic mutations in the Shwachman-Bodian-Diamond syndrome (*SBDS*) gene are responsible for 90% of SDS patients ^[13-16]^. Gene conversion between *SBDS* and its unprocessed pseudogene *SBDSP1* results in two most common functional paralogous sequence variants (PSVs), c.183_184delinsCT and c.258+2T>C, on exon 2^[17]^. These two functional PSVs account for approximately 80% of the *SBDS* disease alleles ^[16, 18-20]^, indicating the significance of accurate variant detection at these two loci in SDS molecular diagnosis.

Disentangling the disease-causing variants through short NGS reads is frequently limited since *SBDS* shares 97% identity with *SBDSP1* ^[21-23]^. Yamada *et al*. reported two SDS patients with compound heterozygous mutations in the *SBDS* gene (c.258+2T>C on one allele and c.183_184delinsCT together with c.201A>G on the other), validated by Sanger sequencing. However, the c.183_184delinsCT and c.201A>G variants were not identified by whole-exome sequencing (WES) in either patient due to mismapped reads, which pointed out the shortcomings of NGS data analysis for SDS diagnosis ^[22]^. Since SDS usually presents with clinical heterogeneity and is thought to be underdiagnosed in the general population ^[12]^, the omittance of pathogenic variants limited by ambiguous read mapping during conventional NGS analysis might be one of the possible causes.

Although the American College of Medical Genetics and Genomics (ACMG) guidance has pointed out that “laboratories must develop a strategy for detecting disease-causing variants within regions with known homology” ^[24]^, to our knowledge, few studies have confronted the analysis of variants arising from gene conversion based on short-read NGS data. In addition, although inaccurate molecular pathogenesis detection of the two most common *SBDS* gene PSVs has been scatteredly reported ^[22]^, a systematic description of the degree of error of standard analysis has seldom been assessed.

In this study, we developed an efficient computational approach named HapICE based on the Expectation-Maximization (EM) algorithm to infer the haplotype consisting of the PSVs for *SBDS* molecular diagnosis and carrier screening through regular short-read NGS data. We retrospectively analyzed the two functional PSVs arising from *SBDS*/*SBDSP1* gene conversion among 47 suspected SDS patients by HapICE. The novel tool’s analysis result was 100% (95% CI: 92.5%-100%) consistent with Sanger sequencing and showed an improved diagnostic rate of 27.7% compared with conventional NGS analysis, highlighting its advantages over standard NGS analysis for the detection of complex variants derived from gene conversion in clinical scenarios. We further compared the variant detection result among the enrolled 47 individuals and 64 available parents between two methods, finding that conventional NGS analysis resulted in 17.0% inaccurate pathogenesis diagnosis, 25.2% false-negative, and 1.8% false-positive variant detections in the overall 111 individuals.

## 2. Methods

### 2.1 Study design and data collection

The outline of the study design is shown in **Figure 1**. The SDS high-risk cohort was retrospectively collected from deidentified individuals submitted to the Children’s Hospital of Fudan University (CHFU) for genetic testing, with suggestions from clinicians and informed consent from parents. The inclusion criteria were patients who were 1) carriers of *SBDS* variants of the two functional PSVs (c.183_184delinsCT and c.258+2T>C) or 2) clinically diagnosed as SDS, and conventional NGS analysis detected one heterozygous variant of either functional PSV. The detailed clinical inclusion criteria included 1) hematologic abnormalities characterized by persistent or intermittent neutropenia and 2) liver dysfunction with persistently elevated serum aminotransferase. Patients were excluded if 1) they were genetically diagnosed by pathogenic/likely pathogenic (P/LP) SNVs/small indels on other exons of *SBDS* or other established SDS-related genes or 2) they had severe infections, chronic viral hepatitis, toxicity and drug-induced liver injury, autoimmune diseases, or hemophagocytic syndrome. The detailed sequencing information is described in the **Supplementary Notes**.

**Figure 1.**
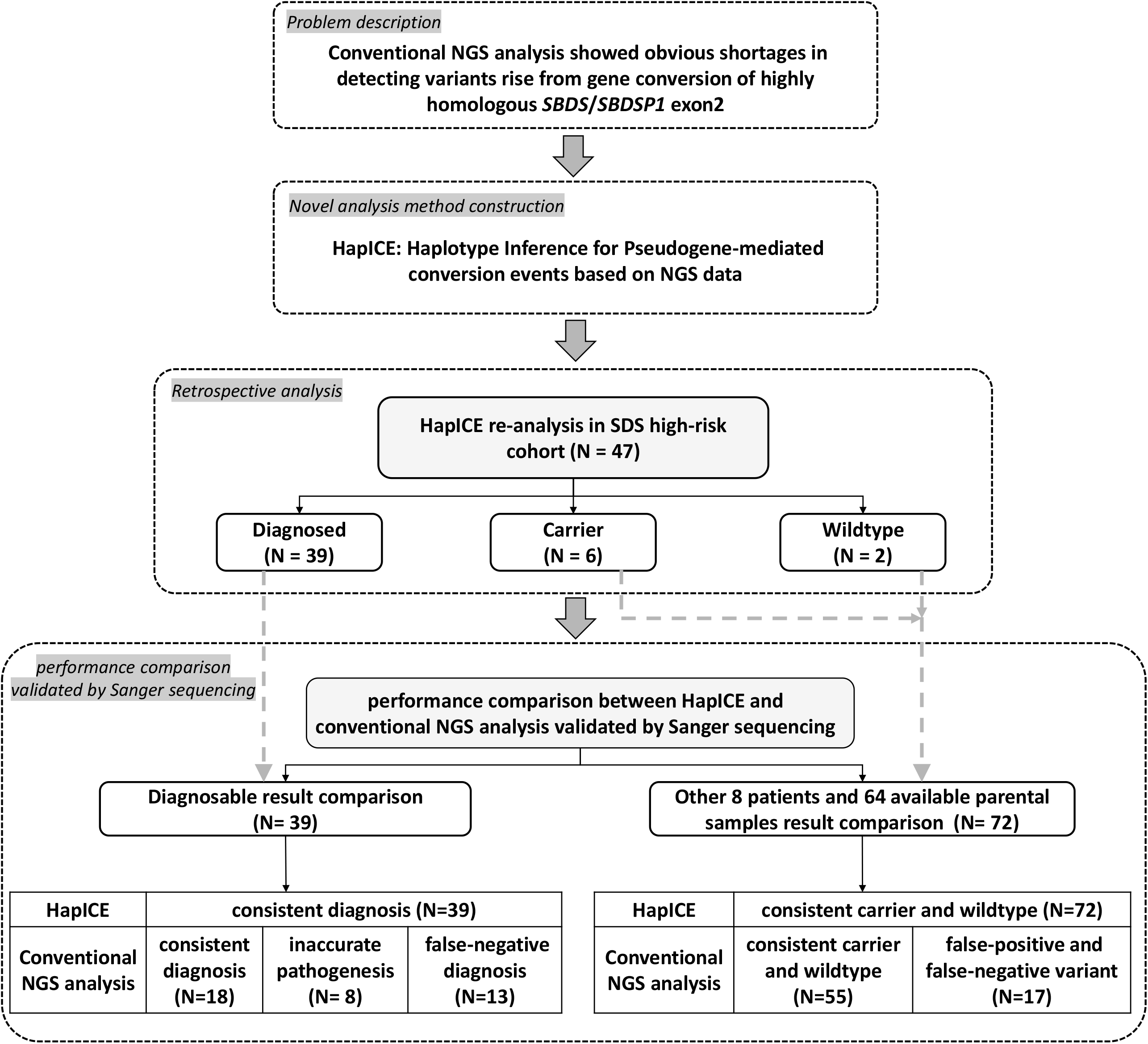
The outline of the study design. We described the potential pitfall of conventional NGS analysis in detecting the most common pathogenic *SBDS* variants and then proposed a novel solution, HapICE, to optimize the variant analysis. We then applied HapICE for retrospective analysis in an SDS high-risk cohort for molecular diagnosis and validated by Sanger sequencing. We further evaluated and compared variant detection performance between HapICE and conventional NGS analysis according to Sanger sequencing results.

### 2.2 Conventional NGS variant calling of SNV/small indels

Variant calling of SNV/small indels from NGS data for conventional genetic diagnosis was performed following the pipeline described in our previously published studies^[1, 25]^. In brief, the sequencing reads were aligned to the GRCh37 (hg19) reference human genome using the Burrows-Wheeler Aligner (0.7.15-r1140)^[26]^. SAMtools (v.1.8)^[27]^ and Picard tools (v.2.20.1, http://broadinstitute.github.io/picard/) were used to sort, merge, and remove duplicated BAM files. SNVs and small indels were detected using the GATK tool and Best Practice Pipelines with default parameters (https://software.broadinstitute.org/gatk/best-practices).

### 2.3 Haplotype inference for pseudogene-mediated conversion events based on short-read NGS data

A computational approach, HapICE (Haplotype Inference for pseudogene-mediated Conversion Events), was established to detect variants created from *SBDS*/*SBDSP1* gene conversion through short sequencing reads. The four main steps include:

#### (1) Prepare the gene-specific combined reference

To identify the consistent and informative regions of the *SBDS*/*SBDSP1* genes, the combined reference and annotation files were generated by the following steps: (i). Generate a combined reference file by *SBDS*/*SBDSP1* gene alignment with MUSCLE ^[28]^ (v3.8.31), in which inconsistent bases between two genes are kept as origins and other consistent bases are marked as “N”; (ii). Generate a transition file that transferred the position on the combined reference to the original genomic position of the *SBDS*/*SBDSP1* genes; (iii). Generate the region annotation file in which the combined reference was classified into the target regions (different bases) and the other regions (consistent bases).

#### (2) Generate the reads-to-region mapping content dataset

To collect the relevant reads that support the inference of potential haplotypes, reads from the BAM file were realigned to the generated combined reference sequence. First, reads that mapped to any position of the *SBDS*/*SBDSP1* genes were extracted by SAMtools ^[29]^. Then, the extracted reads were remapped to the combined reference by BLAT ^[30]^. Finally, the remapping result was transferred to a “reads-to-region mapping content” dataset, in which each row presented an extracted read, each column presented a target region, and each element recorded the exact base information at a given genomic region supported by a specific sequencing read.

#### (3) Parental gene/pseudogene haplotype inference

##### I: Problem statement for haplotype inference

A categorical distribution was used to describe the haplotype information. Assuming there are M possible haplotypes {*h*_1_, *h*_2_, …, *h*_*M*_} covering K target regions, the corresponding probability for haplotypes is θ = {*θ*_1_, *θ*_2_, …, *θ*_*M*_}, with 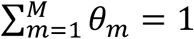. The categorical distribution is recorded as 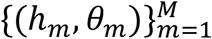. The goal of the algorithm is to estimate the haplotype probability θ.

##### II: Haplotype inference by Expectation-Maximization

The EM algorithm^[31]^ was used to estimate the probability parameters {*θ*_*M*_} for haplotype {*h*_*M*_} with sequencing reads *x*_*n*_ as the observations. We use *δ*(*x, h*) to indicate whether a certain read *x* can support haplotype *h* (at least two intersecting target regions had no conflict). For the total M haplotypes and N reads, a binary matrix *D*_*n*\**m*_ with ensembles {*δ*_*nm*_} was used to denote whether read *n* supported haplotype *m*.

###### The E-step

Suppose the prior distribution for *h*|*θ* is the categorical distribution 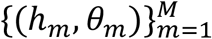. It can be derived that the posterior distribution for *h*|x, *θ* is 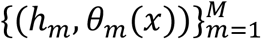, where

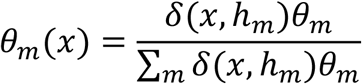

Given parameters *θ*^(*t*)^ at step t and sequencing read data {*x*_*n*_}, the conditional expected log-likelihood Q-function is:

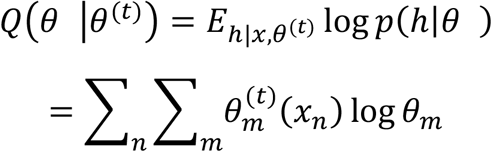

###### The M-step

Maximizing the Q-function

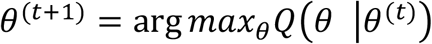

yields

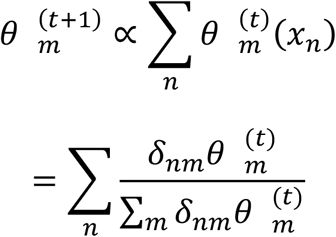

###### Algorithm implementation

Initially, 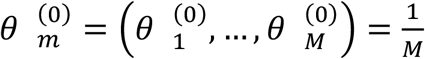. Here, 1/M is not mandatory, and other values also work.

At each time t,

Calculate the posterior distribution matrix 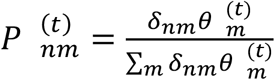

Sum according to haplotypes 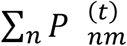 and normalize to obtain 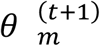

The iteration step will stop if convergence occurs 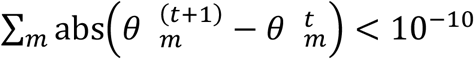.

##### III: Reduce calculation complexity by a pruning and recursive strategy

###### Problem statement

each region *T*_*k*_ contains the collection set of all possible sequence contexts in this region. Theoretically, the haplotype’s state space is the Cartesian production of contexts of the K regions. Nevertheless, the probability values for most of the possible haplotypes are zero. Thus, we applied a recursive pruning strategy to reduce the calculation complexity.

###### Algorithm implementation

For k=1, directly perform the trivial estimation.

For k>1, augment the state space from k-1 to k, i.e., haplotypes for k is the sequence concatenated from all remaining haplotypes from k-1 with all possible sequence context at region k. Do the EM step first and only reserve haplotypes with the highest probabilities (e.g., top 3 haplotypes) or only reserve haplotypes with probabilities higher than 0.01.

Output the final haplotypes with the corresponding probability when k=K.

#### (4): Result visualization

The proportion of gene recombination events between two neighboring informative bases, haplotype supportive read information, and the final inferred haplotypes of the parental gene/pseudogene pairs are displayed.

### 2.4 Sanger validation

Sanger sequencing was adopted to unambiguously study the mutational profile of *SBDS* and *SBDSP1* using a well-established protocol developed in our laboratory. The protocol was based on long-range PCR amplification with *SBDS* exon 2 allele-specific primers (forward: 5’-CTGCACCCCACCCCACCC-3’, reverse: 5’-TAAAAAATGAGTAACTGGATGGAG-3’), followed by DNA sequencing of smaller fragments (forward: 5’-AAAGAAAACTGCCCTCTACAC-3’, reverse: 5’-TCACATTATTGCTTGGTTAGTC-3’).

## 3. Results

### 3.1 NGS sequence alignment at *SBDS*/*SBDSP1* and possible interferences when analyzing gene conversion events

Exon 2 of *SBDS* and *SBDSP1* differed by only seven bases, and the relatively short sequencing reads frequently fell into homologous pitfalls from ambiguous alignment, resulting in incorrect variant calling, especially when variants arisen from gene conversion events (**Figure 2**). For example, we confronted a patient (Case 3) with compound heterozygotes of two pathogenic *SBDS* alleles (the c.258+2T>C allele and another allele comprised of c.183_184delinsCT and c.201A>G) determined by trio Sanger sequencing. However, only the c.258+2T>C variant was correctly detected by conventional NGS analysis (**Figure 2B**). A manual review found that nearly all of the NGS reads containing the two *SBDS* variants, c.183_184delinsCT and c.201A>G, were incorrectly aligned to the *SBDSP1* gene and mistakenly regarded as wild-type reads derived from *SBDSP1*. At the same time, n.424 and n.533+10 of the *SBDSP1* gene were both mistakenly called heterozygous variants (**Figure 2B** and **Figure 2C**). The ambiguous mapping resulted in false-negative variant callings for *SBDS* and false-positive callings for *SBDSP1* in regular NGS data analysis.

**Figure 2.**
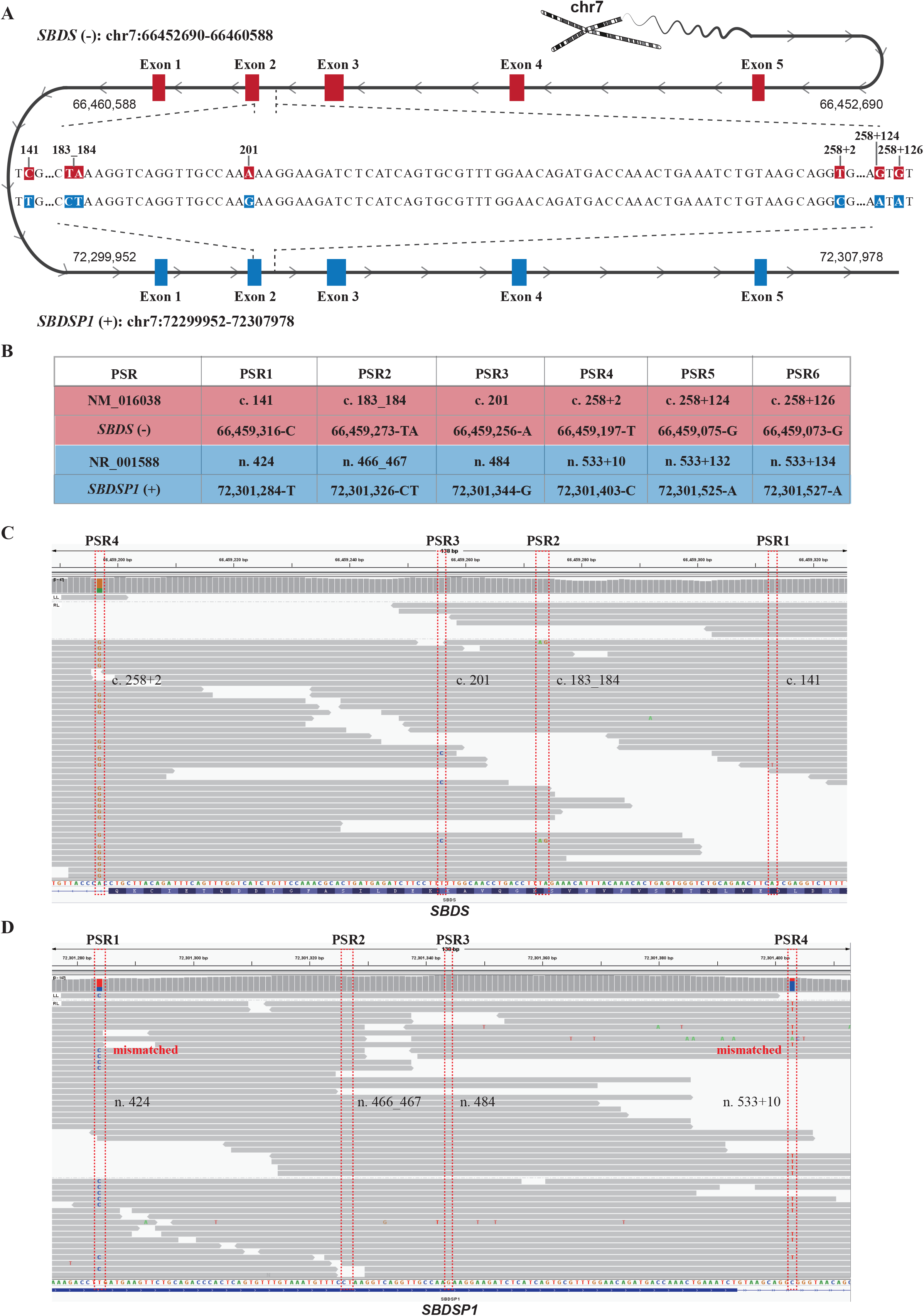
Schema of gene conversion between *SBDS* and its pseudogene *SBDSP1* and the NGS sequence alignment surrounding the functional PSVs. (**A**). There are only six PSRs (paralogous sequence regions) that are different between the exon 2 of *SBDS* (reverse strand) and the *SBDSP1* (forward strand) gene. (**B**) The chromosome location, reference base, and transcript location of the six PSRs between *SBDS* exon 2 and *SBDSP1*. Among them, only two functional *SBDS* PSVs (paralogous sequence variants), c.258+2T>C and c.183_184delinsCT, would affect the protein-coding and are often used as the *SBDS*/*SBDSP1* gene conversion events’ markers. (**C**) and (**D**) show the NGS sequence pileups of read pairs (2*150) in a sample with two wild-type copies of *SBDSP1* and one copy of *SBDS* exon 2 with a heterozygous c.258+2T>C variant and another *SBDS* allele with both heterozygous c.183_184delinsCT and c.201A>G variants confirmed by trio Sanger sequencing. On the *SBDS* locus **(C)**, the c.183_184delinsCT and the c.201A>G variants were missed due to the ambiguously mapped reads. Meanwhile, the false-positive variants n.424T>C and n.533+10C>T were called on the *SBDSP1* locus (**D**).

### 3.2 Haplotype inference for *SBDS*/*SBDSP1* gene conversion based on NGS data

To solve the problem of inaccurate variant detection caused by the interference of highly homologous sequences, we developed an automatic tool, HapICE, to detect variants arising from gene conversion. We took the *SBDS* c.183_184delinsCT and c.258+2T>C variants that were generated from *SBDS*/*SBDSP1* gene conversion as an application example (**Figure 3**).

**Figure 3:**
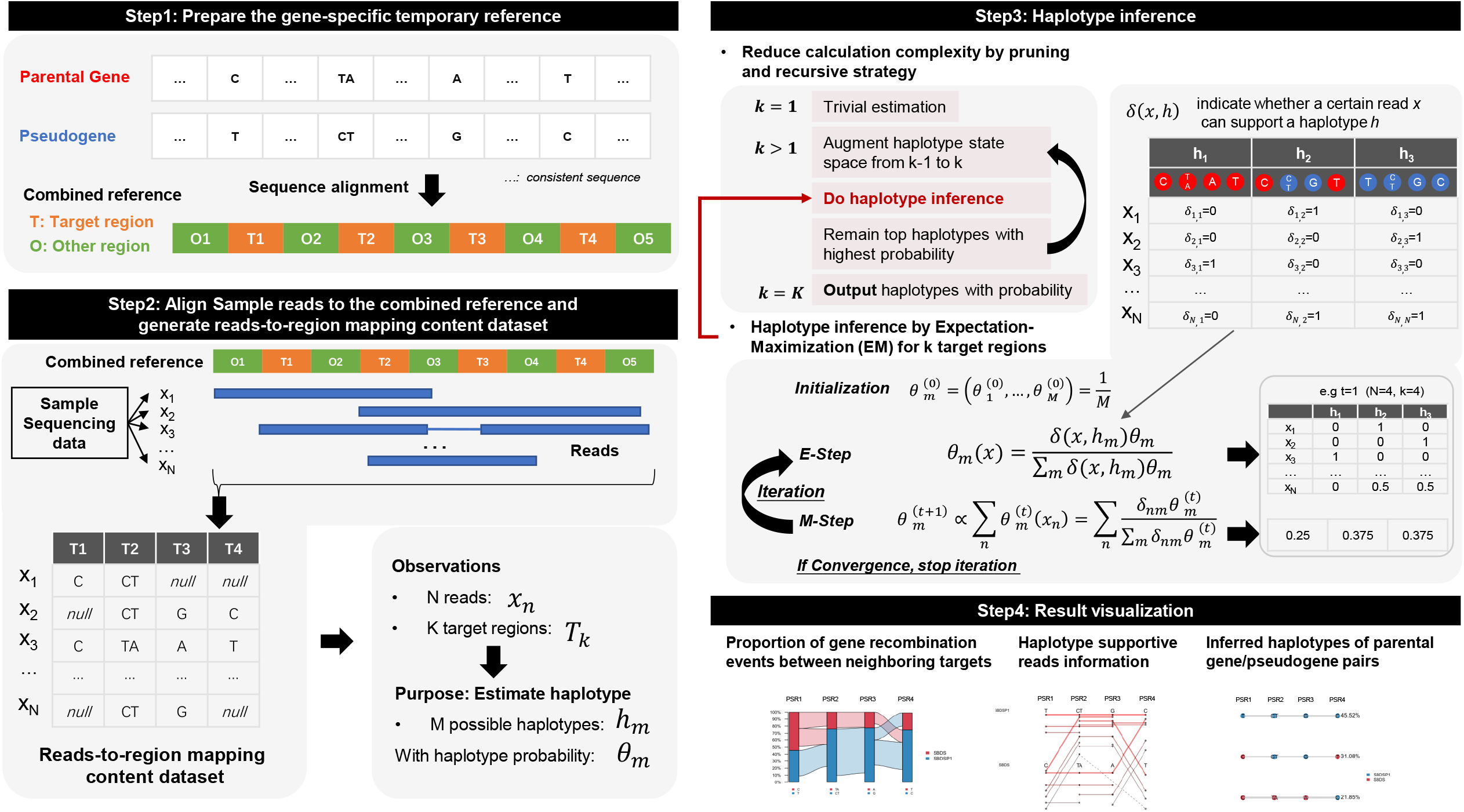
A novel computational algorithm HapICE for *SBDS*/*SBDSP1* gene conversion analysis using next-generation sequencing data. HapICE involves four main steps for *SBDS*/*SBDSP1* gene conversion analysis. Step1: Prepare the gene-specific combined reference. Genome sequences from parental and pseudogene are aligned and marked into *target region* (where bases in parental gene/pseudogene genes were different, often PSVs) and *other region* (where bases were consistent). Step2: Align reads to the combined reference, and generate reads-to-region mapping content dataset. Sequencing reads are aligned to the combined reference and the exact base information located at the target region is recorded to generate the dataset. This dataset describes the mapping observation and is used to estimate haplotypes with probabilities. Step3: Haplotype inference. A pruning and recursive strategy is used to reduce calculation complexity and for each k (k>1), the haplotype state space is augmented from k-1, and the haplotypes with the highest probabilities inferred from the Expectation-Maximization (EM) step are reserved for the next step. For the EM step for k target region, the probability for each candidate haplotype was initialized, expected (E-Step) and maximized (M-Step) until convergence. Step4: Result visualization. Three visualization functions are provided to show the haplotype structures with probabilities, detailed recombination events, and supportive reads information.

Generally, since the read-mapping result in homologous regions is largely determined by PSVs, the PSV loci between *SBDS* and *SBDSP1* can be used as anchor points to guide the short read mapping process. By inferring the haplotype block of these PSVs and calculating their corresponding proportions, HapICE can detect the variants that arise from gene conversion events and help make molecular diagnoses. Specifically, we aligned the genomic sequence of the *SBDS*/*SBDSP1* gene and generated a new combined reference to identify the consistent and informative regions of the genes. Then, we realigned the sequencing reads (from FastQ or BAM) to the combined reference sequence to collect supportive reads for the potential haplotypes of PSVs. Next, we performed EM algorithm by considering the haplotype composed of PSVs as the latent variable to infer the conversion haplotypes and reduce the calculation complexity by a pruning and recursive strategy. The haplotype inference result was eventually visualized in multiple aspects (**Figure 3**). The HapICE package is open source and available online at https://github.com/SherryDong/HapICE.

### 3.3 Retrospective analysis of the SDS high-risk cohort

After novel tool construction, we performed a retrospective analysis of c.183_184 and c.258+2 loci of the *SBDS* gene in an SDS high-risk cohort. Altogether, 47 individuals from 46 unrelated families met the inclusion criteria, including 29 boys and 18 girls aged from newborn to 6 years old. Fourteen and 33 patients underwent clinical-exome sequencing (CES) and WES, respectively, with an average sequencing coverage of 199X and 103X.

Altogether, the HapICE reanalysis results showed that 39 (83.0%) patients obtained diagnosable *SBDS* haplotypes, and the other 6 (12.8%) and 2 (4.3%) individuals were carriers and wild-type at these two functional PSV loci, respectively (**Supplementary Table 1**). For the 39 patients with diagnosable *SBDS* haplotypes, 31 (79.5%) samples had both heterozygous variants, 2 (5.1%) harbored a homozygous variant of c.258+2T>C, and the other 6 (15.4%) had an allele of c.258+2T>C together with another allele of c.[183_184delinsCT;258+2T>C]. We performed Sanger sequencing on all of the enrolled patients and found that HapICE achieved 100% (95% CI: 92.5%-100%) consistent variant detection results compared with the orthogonal method, demonstrating HapICE’s ability to accurately detect functional PSVs arising from gene conversion events. Through HapICE reanalysis, a diagnostic rate of 83.0% (39/47) was achieved in this SDS high-risk cohort.

Moreover, HapICE was able to determine the phasing of the PSV haplotypes (*in cis*/*trans*) through proband-only analysis, while parental validation was necessary for Sanger sequencing to confirm compound heterozygous variants. Among the 31 diagnosable patients who had both heterozygous PSVs, 24 were available for further parental Sanger validation. HapICE haplotype results showed that the two functional PSVs were all *in trans*, which is consistent with the parental Sanger sequencing.

### 3.4 Comparison between the HapICE result and conventional NGS analysis

Since all of the variant detection results of HapICE were confirmed by Sanger sequencing, we then compared the HapICE result with conventional NGS analysis among the high-risk SDS cohort and evaluated their variant detection performances.

In conventional NGS analysis, 26 (55.3%) samples had a potential molecular diagnosis, including 10 with a homozygous c.258+2T>C variant and 16 harboring both heterozygous functional PSVs. Another 21 individuals were identified as carriers, including three with a heterozygous c.183_184delinsCT variant and 18 with a heterozygous c.258+2T>C variant (**Table 1**).

**Table 1.**
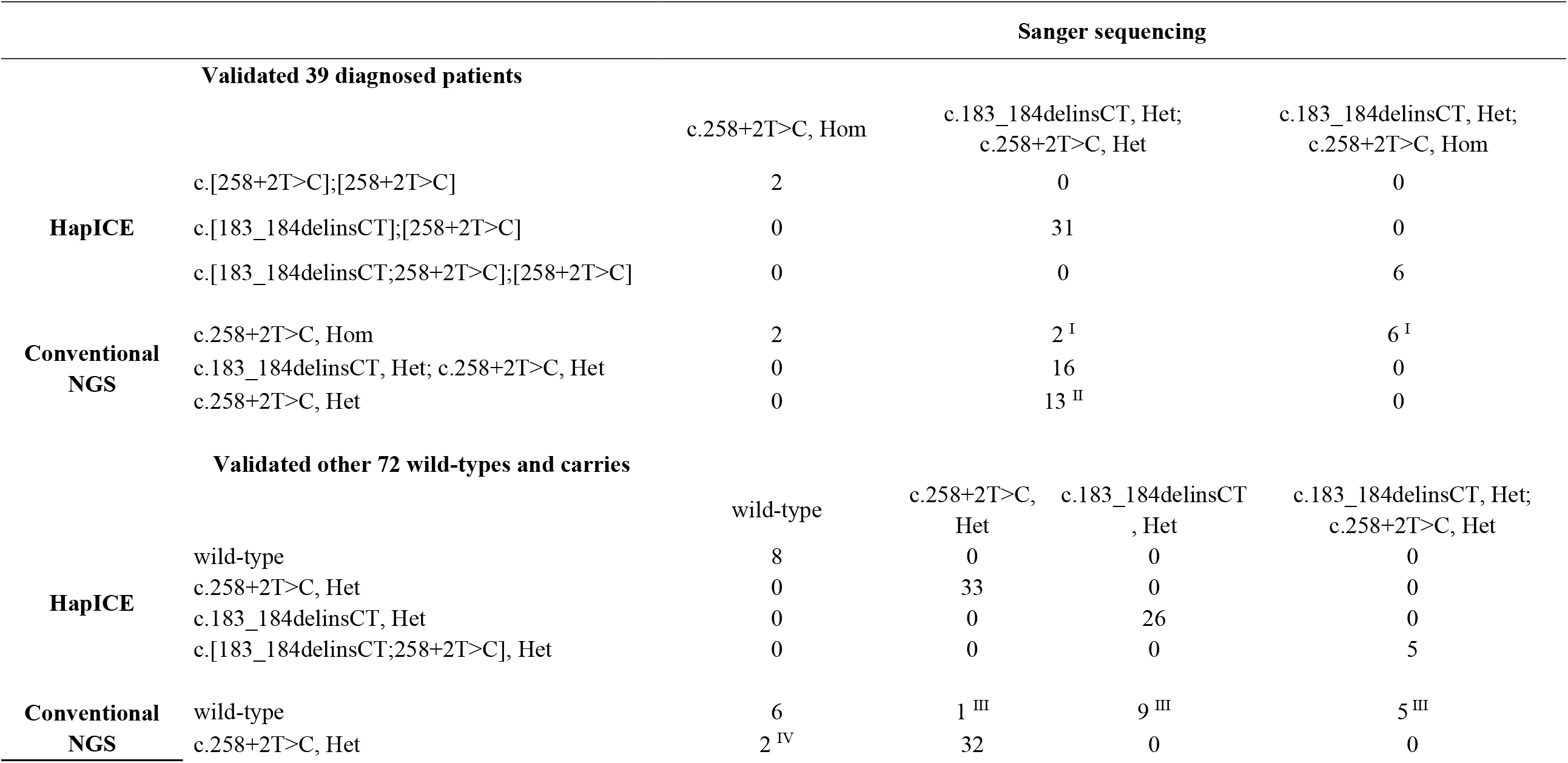
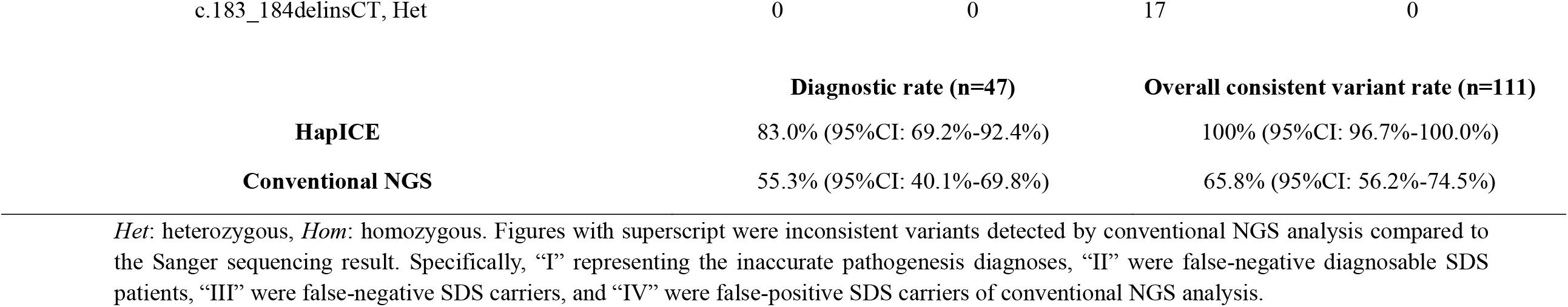
Comparison of HapICE and conventional NGS data analysis for *SBDS* exon2 functional PSVs validated by Sanger sequencing.

|  |  | Sanger sequencing |  |  |  |
| --- | --- | --- | --- | --- | --- |
| Validated 39 diagnosed patients |  |  |  |  |  |
|  |  | c.258+2T>C, Hom | c.183_184delinsCT, Het;<br>c.258+2T>C, Het |  | c.183_184delinsCT, Het;<br>c.258+2T>C, Hom |
| HapICE | c.[258+2T>C];[258+2T>C] | 2 | 0 |  | 0 |
|  | c.[183_184delinsCT];[258+2T>C] | 0 | 31 |  | 0 |
|  | c.[183_184delinsCT;258+2T>C];[258+2T>C] | 0 | 0 |  | 6 |
| Conventional<br>NGS | c.258+2T>C, Hom | 2 | 2 <sup>I</sup> |  | 6 <sup>I</sup> |
|  | c.183_184delinsCT, Het; c.258+2T>C, Het | 0 | 16 |  | 0 |
|  | c.258+2T>C, Het | 0 | 13 <sup>II</sup> |  | 0 |
| Validated other 72 wild-types and carries |  |  |  |  |  |
|  |  | wild-type | c.258+2T>C,<br>Het | c.183_184delinsCT<br>, Het | c.183_184delinsCT, Het;<br>c.258+2T>C, Het |
| HapICE | wild-type | 8 | 0 | 0 | 0 |
|  | c.258+2T>C, Het | 0 | 33 | 0 | 0 |
|  | c.183_184delinsCT, Het | 0 | 0 | 26 | 0 |
|  | c.[183_184delinsCT;258+2T>C], Het | 0 | 0 | 0 | 5 |
| Conventional<br>NGS | wild-type | 6 | 1 <sup>III</sup> | 9 <sup>III</sup> | 5 <sup>III</sup> |
|  | c.258+2T>C, Het | 2 <sup>IV</sup> | 32 | 0 | 0 |

#### 3.4.1 Diagnosable result comparison

##### (1) Consistent diagnosis determined by HapICE and conventional NGS analysis

All 26 patients with a diagnosable result by conventional NGS analysis were fully covered by HapICE diagnosable patients. However, only 18 (69.2%) patients had consistent genetic variant conclusions by both methods, including 2 who had a homozygous c.258+2T>C variant and 16 who harbored both heterozygous functional PSVs (**Table 1**).

##### (2) Inaccurate pathogenesis diagnosis by conventional NGS analysis

Apart from the consistent diagnosis cases, the molecular pathogenesis of 8 samples (30.8%, 8/26) was inaccurately reported by conventional NGS analysis. Specifically, two individuals were mistakenly called the *SBDS* homozygous c.258+2T>C variant but were compound heterozygous at c.183_184delinsCT and c.258+2T>C. The other six individuals were validated to simultaneously have the homozygous c.258+2T>C and heterozygous c.183_184delinsCT variants, while conventional NGS analysis only detected the homozygous c.258+2T>C variant.

##### (3) False-negative missed by conventional NGS analysis

False-negative results were also detected by conventional NGS analysis during SDS molecular diagnosis. For the validated diagnosable samples, 13 individuals with both heterozygous c.183_184delinsCT and c.258+2T>C variants were missed by conventional NGS analysis. These false-negative cases accounted for 41.9% (13/31) of all diagnosable individuals with both heterozygous functional PSVs, improving the diagnostic rate by 27.7% (13/47) through HapICE compared with conventional NGS analysis. All 13 cases identified only a heterozygous c.258+2T>C variant through conventional analysis. A further manual review revealed that the interference resulted from the c.141C>T and/or c.201A>G nonfunctional PSVs. When these two nonfunctional PSVs came with c.183_184delinsCT *in cis*, sequencing reads covering both variants were incorrectly mapped to *SBDSP1* in conventional analysis and thus missed c.183_184delinsCT (**Figure 2C** and **D**).

These comparison results showed that when both variants appeared on the two functional loci of *SBDS*, 56.8% (21/37) of the c.183_184delinsCT variant was missed by conventional NGS analysis. In contrast, HapICE reanalysis could result in an improved diagnostic rate of 33.3% (13/39) and a more precise pathogenesis conclusion rate of 30.8% (8/26) compared with conventional NGS analysis, demonstrating its application potential in the clinical molecular diagnosis scenario.

#### 3.4.2 Other patients and available parental samples analysis result comparison

We further investigated the variant detection results of HapICE and conventional NGS analysis at the two functional PSVs among the remaining 8 undiagnosed patients and 64 available parental samples of the cohort. Variant detection conclusions of the two methods were compared based on Sanger sequencing.

For these 72 individuals, HapICE showed that 64 and 8 were carriers and wild-type, respectively, while 51 carriers and 21 wild-type individuals were detected by conventional NGS analysis. Sanger sequencing was again 100% (95% CI: 95.0%-100%) consistent with HapICE, while conventional NGS analysis made incorrect variant calling in 17 (23.6%) samples (**Table 1**).

##### (1) Consistently identified carrier and wild-type individuals

Altogether, 55 individuals were consistently identified at these two functional PSVs by HapICE and conventional NGS analysis, including 49 carriers (6 patients and 43 parental samples) and 6 wild-type individuals (all parental samples).

##### (2) False-positive and false-negative carriers detected by conventional NGS analysis

The inconsistently identified individuals reflected the false-positive and false-negative carrier callings of conventional NGS analysis at the c.183_184 and c.258 loci. Specifically, among 17 samples with inconsistent results, two were false-positively called a heterozygous c.258+2T>C variant by conventional NGS analysis. However, both HapICE and Sanger sequencing showed wild-type *SBDS* and HapICE reported *in cis* n.484G>A and n.466_467delinsTA in *SBDSP1*. These results showed that when both variants appeared on n.466_467 and n.484 of the *SBDSP1* gene, conventional NGS analysis would prefer to call a false-positive c.258+2T>C variant on *SBDS*. The other 15 parents were false-negatively identified as wild-type by conventional NGS analysis, while validated to include one with heterozygous c.258+2T>C, nine with heterozygous c.183_184delinsCT, and five with two functional PSVs *in cis*.

The overall comparison of variant detection of the two functional PSVs on *SBDS* between HapICE and conventional NGS analysis showed that HapICE achieved an improved diagnostic rate of 27.7% in the SDS high-risk cohort (n=47) and a consistent variant detection rate of 100% (95% CI: 96.7–100%) among all validated individuals (n=111). Conventional NGS analysis only showed a consistent rate of 65.8% (95% CI: 56.2%-74.5%), resulting in both false-negative and false-positive conclusions in SDS molecular pathogenesis analysis.

## Discussion

Shwachman-Diamond syndrome, primarily resulting from parental/pseudogene (*SBDS/SBDSP1*) conversion events, is a representative issue in NGS-based molecular diagnosis scenarios. Generally, when SDS is highly suspected based on typical clinical features, PCR sequencing with specific primer pairs can be applied to detect variants selectively within the *SBDS* locus. However, the disease is thought to be underdiagnosed because of ambiguous clinical presentations ^[12]^. In a clinical scenario where patients’ disease-related phenotypes are mild and atypical, especially during infancy, exome sequencing that can screen for the entire gene set is commonly applied in pathophysiology examinations. Thus, it is of extreme importance to accurately detect pathogenic *SBDS* variants resulting from gene conversion based on regular exome sequencing data. However, to our knowledge, few attempts have been made to provide a corresponding solution in previous studies, let alone systematic evaluation of the variant detection errors standard analysis would make, yet these works are essential for a better understanding of SDS.

In this study, we presented a novel tool, HapICE, to infer the haplotype consisting of PSVs for molecular diagnosis and carrier screening through conventional NGS data. We applied HapICE for reanalysis of two *SBDS* functional PSVs in an SDS high-risk cohort involving 47 Chinese pediatric patients. HapICE achieved a completely consistent result with Sanger sequencing and reached a diagnostic rate of 83.0%. Compared with conventional NGS analysis, HapICE showed an improved diagnostic rate of 27.7% and an amended variant calling rate of 17.0% in this cohort. In addition to precise variant detection, HapICE could identify whether two heterozygous functional PSVs were *in cis* or *trans* by inferring the haplotype blocks from proband-only data. These results highlight HapICE as an efficient assay for the diagnosis of SDS in place of conventional NGS analysis.

Moreover, we systematically analyzed the variant detection features of c.258+2 and c.183_184 through conventional NGS analysis. We found that conventional NGS analysis for *SBDS* gene exon 2 functional PSV callings would be frequently influenced by *SBDSP1* or other *SBDS* nonfunctional PSVs, resulting in 25.2% false-negative and 1.8% false-positive variant detections, which may worsen the SDS underdiagnosis situation. The complexities of pathological diagnosis and the indispensable treatment of SDS patients highlight the need for efficient HapICE to be adopted in a large NGS diagnosis and carrier-testing platform to complement routine exome sequencing data analysis.

In summary, as exome sequencing has become a preferred tool for molecular diagnosis, it is critical to involve gene conversion events that affect highly homologous regions such as *SBDS*/*SBDSP1* for more reliable variant detection. Although long-read sequencing technologies can largely overcome the inaccurate alignment problem of homologous reads, these platforms still either generate many sequencing errors or are too costly to be implemented in diagnostic laboratories. In this study, we showed that conventional NGS analysis can lead to the incomplete delineation of molecular pathogenesis and described the extent of the mistakes it will make in analyzing the two common functional PSVs of *SBDS*. We proposed a novel tool to solve the difficulty based on conventional short-read NGS data, which showed fantastic variant detection performance in an SDS cohort. Moreover, we applied HapICE to our in-house spinal muscular atrophy (SMA, MIM#253300) patients to detect the functional PSV of *SMN1* c.840C>T that results from highly homologous *SMN1* and *SMN2* gene conversion events ^[5]^. The results were confirmed by Sanger/multiplex ligation-dependent probe amplification (MLPA) (data not shown), demonstrating HapICE’s potential to be further optimized and promoted to a wider range of clinical scenarios.

However, there are still some limitations in this study that can be further improved. The present study only focused on the two most common variants in the *SBDS* gene exon 2, and other pathogenic variants were not thoroughly explored. In addition, previous studies reported that structural variation (SV) in *SBDS* was associated with SDS. Structural alterations, such as large deletions, duplications, insertions, or inversions, are anticipated due to the peculiar genomic architecture of the *SBDS*/*SBDSP1* loci ^[32]^. When the detected SNVs are insufficient to draw diagnosable conclusions in highly suspected clinical patients, the presence of SVs should be considered. However, HapICE did not address these situations and mainly focused on functional PSVs, which should be further improved for comprehensive variant analysis.

## Data Availability

The data that support the findings of this study are either included in the article (or in its supplementary files) or available from the corresponding author on reasonable request.

## Data availability statement

The data that support the findings of this study are either included in the article (or in its supplementary files) or available from the corresponding author on reasonable request. The data are not publicly available due to privacy or ethical restrictions.

## Acknowledgment

We thank the patients and their families, and referring physicians for their contribution to this study. We would like to thank Mr. W.B. Sheng for providing mathematical algorithm suggestions. This work was funded by the National Key Research and Development Program (2018YFC0116903), Shanghai Municipal Science and Technology Major Project (2017SHZDZX01), the fellowship of China Postdoctoral Science Foundation (2020M681176), Shanghai Key Laboratory of Birth Defects (13DZ2260600), Shanghai Municipal Science and Technology Major Project (20Z11900600) and Clinical Skill and Innovation program of Shanghai Shenkang Hospital Development Center (SHDC12020CR6028-002).

## Author Contributions

Conceptualization: Y.L., X.D., B.L., and W.Z.; Data curation: X.P., W.L., F.X., and L.Y.; Formal analysis: B.L., X.P., Y.L., X.D., and B.W.; Software & Visualization: X.D., B.L., and Y.L.; Validation experiments: G.L.; Writing-original draft: B.L., X.P.; Writing-review & editing: X.D., Y.L., Y.W., and H.W. All authors read and approved the final manuscript. The authors wish it to be known that, in their opinion, X.P. and X.D. should be regarded as joint First Authors.

## Ethics Declaration

This study was approved by the ethics committees of Children’s Hospital of Fudan University (2015–130). Counseling was performed by physicians before testing. Informed consent was obtained from the parents of each patient.

