## Supplementary Note for "Overcoming the pitfalls of NGS-based molecular diagnosis of Shwachman-Diamond syndrome"

### Next-generation sequencing of the cohort

Clinical exome sequencing (CES) or whole-exome sequencing (WES) was performed between May 2016 and February 2021 following the protocol described in our previous studies[1, 2]. In brief, DNA was extracted from peripheral blood specimens according to the manufacturer's instructions using the Thermo Fisher Scientific (Shanghai, China) KingFisher LabServ kit. Genomic DNA was enriched using the Agilent ClearSeq Inherited Disease Kit for CES; and the Agilent SureSelect All Exon Human V5 Kit (Santa Clara, CA, USA), IDT xGen Exome Research Panel V2 Kit (Coralville, Iowa), or NanoWES Human Exome Kit (Beijing, China) for WES. DNA fragments were ligated with adaptors to generate two paired-end DNA libraries with an average insert size of 500 bp. After enrichment via the polymerase chain reaction, the DNA libraries were sequenced on the Illumina HiSeq2000/2500 Platform or Illumina NovaSeq 6000(San Diego, CA, USA) to yield 150-bp paired-end sequencing reads.
